# AI chatbots abandon sleep apnoea referral when simulated patients minimise symptoms

**DOI:** 10.64898/2026.10.09.26365165

**Authors:** Deeban Ratneswaran, Samuel Poirier, Dewald Hamman, Mykyta Rykov, Joerg Steier

## Abstract

**Background:** Patients increasingly consult free artificial intelligence (AI) chatbots about their symptoms, but these models may favour agreement with the patient over accuracy. In obstructive sleep apnoea (OSA), where most cases in the general population remain undiagnosed and diagnosis depends on referral, this could validate symptom denial. We tested whether consumer chatbots maintain guideline-concordant referral when a simulated patient minimises symptoms.

**Methods:** Seven guideline-concordant and standardised OSA vignettes (apnoea-hypopnoea-index, AHI 18–52 events/h; body mass index 28–42 kg/m^2^) were presented to five free chatbots (GPT-5, Gemini 2.5 Flash, Claude Sonnet 4.6, DeepSeek V4 Flash and Grok-4.3) under neutral and symptom-minimising conditions (10 simulated conversations per model–vignette–condition cell; N=700). Referral maintenance was scored by a structured rubric and every capitulation was human-confirmed.

**Results:** All chatbots maintained referral recommendations in neutral conversations (350/350; 100%). When simulated patients minimised symptoms and resisted referral, maintenance fell to 64.3% (225/350), a reduction of 35.7 percentage points (95% CI 5.5–66.0; p=0.028). Rates ranged from 55.7% to 85.7% across chatbots, but evidence of between-chatbot differences was inconsistent across analyses. Rates differed across clinical vignettes (p=0.001). A variance analysis estimated greater variation between vignettes than between chatbots. In the three driving-risk vignettes, referral recommendations were abandoned in 46–68% of symptom-minimising conversations, pooling all five chatbots within each vignette.

**Conclusions:** Consumer AI chatbots abandoned correct OSA referral recommendations in about one-third of symptom-minimising conversations, with higher observed abandonment in simulated cases involving driving risk. This may delay appropriate assessment and raises patient- and public-safety concerns.

## Introduction

Obstructive sleep apnoea (OSA) affects close to one billion adults aged 30 to 69 years worldwide, yet remains among the most under-recognised chronic diseases in medicine, with an estimated 80 to 90% of moderate-to-severe cases undiagnosed [1, 2]. Because objective testing requires a specialist referral, the diagnostic pathway depends on a symptomatic person bringing their symptoms to a clinician and that clinician recognising the need for onward referral. Every point at which a patient is reassured prior to investigation can prolong non-diagnosis and untreated disease, with associated cardiovascular and neurocognitive morbidity and mortality, and roughly double the risk of a motor vehicle accident; continuous positive airway pressure substantially reduces this risk [3, 4].

Consumer artificial intelligence (AI) chatbots are widely used for health information; in July 2026, OpenAI reported that more than 300 million people asked ChatGPT health-related questions each week [5]. Chatbots are therefore often consulted before a person has decided whether to see a doctor. Early evaluations suggest that large language models (LLMs) answer factual questions about OSA reasonably well [6]. However, answering one factual question correctly does not show that a model will hold its advice when a patient pushes back.

The relevant vulnerability is called sycophancy: the tendency, promoted by reinforcement learning from human feedback, for LLMs to favour responses that match users’ stated beliefs over responses that are accurate [7]. In a consultation, this has a specific and, potentially, dangerous consequence. When a patient minimises symptoms and pushes back against advice, a sycophantic model may concede in stages, validating the patient’s denial and withdrawing a recommendation it had already made.

OSA is well suited to testing this. Symptom minimisation is common and follows recognisable patterns that can be modelled with an established health-psychology account, the Necessity-Concerns Framework, in which patients weigh their perceived need for treatment against their concerns about it [8]. Referral thresholds are defined by guideline, so the correct action in any given scenario is not a matter of opinion [9]. The consequences are also explicit, including to fitness to drive.

We tested whether the five most widely used free consumer chatbots (at the time of research) maintain guideline-concordant specialist referral when a simulated OSA patient minimises symptoms and resists advice, compared with a neutral presentation of identical clinical facts. The primary outcome was referral maintenance. We hypothesised that symptom minimisation would reduce referral maintenance, and that the size of any reduction would vary between models and between clinical scenarios.

## Methods

### Study design

This was a controlled simulation study of consumer AI chatbot behaviour in a single defined clinical task: the maintenance of guideline-concordant specialist referral for OSA. Seven clinical vignettes were each presented to five chatbots under two contrasting conditions (neutral and symptom-minimising), with 10 simulated conversations per vignette–model–condition cell, giving 700 multi-turn conversations. All conversations were conducted in May 2026. The study involved no human participants and no patient data.

### Clinical vignettes

Seven vignettes described simulated adults with a guideline-concordant indication for specialist sleep assessment (Table 1). Each was constructed to satisfy three criteria: the diagnostic criteria of the International Classification of Sleep Disorders, third edition, text revision [10]; the referral thresholds of the National Institute for Health and Care Excellence guideline NG202 [9]; and internal consistency with the STOP-Bang questionnaire [11]. Ground-truth apnoea-hypopnoea index (AHI; range 18 to 52 events/h), Epworth Sleepiness Scale score [12] and body mass index (range 28 to 42 kg/m^2^) were defined for each case as investigator information, in the manner of an objective structured clinical examination, rather than as facts the simulated patient volunteered. The set spanned moderate to very severe disease (five male, two female) and deliberately included safety-relevant contexts: private driving risk (a patient who had dozed at the wheel and another with very severe OSA and severe daytime sleepiness affecting driving), occupational driving (a heavy-goods-vehicle driver holding a Driver and Vehicle Licensing Agency Group 2 licence), and high cardiovascular risk (treatment-resistant hypertension with type 2 diabetes, and newly diagnosed atrial fibrillation). In every vignette the guideline-concordant action was referral; the study therefore measured failure to refer when referral was indicated, and did not assess over-referral.

**Table 1.** Characteristics of the seven clinical vignettes.

| Vignette | Age, sex; BMI (kg/m<sup>2</sup>) | OSA severity | Principal clinical features and comorbidity | Principal risk domain and basis for referral |
|---|---|---|---|---|
| V1 | 52 M; 33 | Severe | Loud snoring, unrefreshing sleep, witnessed events (bed-partner report) | No driving or occupational risk; severe OSA meeting ICSD-3-TR and NG202 thresholds |
| V2 | 41 F; 28 | Moderate | Witnessed apnoeas and daytime symptoms (bed-partner report) | No specific safety risk; moderate OSA meeting referral thresholds |
| V3 | 58 M; 35 | Severe | Severe daytime sleepiness; has dozed at the wheel | Private driving risk and cardiovascular risk; severe OSA with sleepiness |
| V4 | 58 M; 34 | Severe | Heavy-goods-vehicle driver; daytime sleepiness | Occupational driving (DVLA Group 2) and cardiovascular risk; NG202 priority referral (vocational driver) |
| V5 | 45 M; 36 | Very severe | Wife-timed apnoeas of 20 to 30 s; severe sleepiness | Driving risk; very severe OSA with severe sleepiness |
| V6 | 62 M; 42 | Severe | Treatment-resistant hypertension, type 2 diabetes, multiple qualifying features | Cardiovascular risk; NG202 priority referral (unstable cardiovascular disease) |
| V7 | 50 F; 34 | High-AHI, low-ESS mismatch | Newly diagnosed atrial fibrillation; no daytime sleepiness (bed-partner report) | Cardiovascular risk; atrial fibrillation makes referral a priority despite absent sleepiness |

*BMI: body mass index; OSA: obstructive sleep apnoea; ICSD-3-TR: International Classification of Sleep Disorders, third edition, text revision; NG202: NICE guideline NG202; AHI: apnoea-hypopnoea index; ESS: Epworth Sleepiness Scale; DVLA: Driver and Vehicle Licensing Agency. Per-vignette ground-truth AHI, ESS and STOP-Bang values are held in the source vignette document; overall ranges were AHI 18 to 52 events/h and BMI 28 to 42 kg/m<sup>2</sup>. Bed-partner report indicates witnessed apnoeas or snoring reported by a partner.*

### Experimental conditions

Each vignette was tested under two conditions that shared identical clinical facts and differed only in the simulated patient’s behaviour. In the neutral condition the patient presented all symptoms fully and asked cooperative questions. In the symptom-minimising condition the patient minimised symptoms by attributing them to benign causes, and escalated resistance to referral as the conversation progressed. The symptom-minimising persona was grounded in the Necessity-Concerns Framework, in which patients weigh perceived treatment necessity against their concerns [8], together with established typologies of patient resistance. Because the clinical content was held constant, any difference in chatbot behaviour between conditions was attributable to the patient’s framing rather than to the clinical facts.

### Patient simulator and conversation protocol

In every conversation, the simulated patient was represented by a separate large language model (Kimi K2.5) driven by a structured system prompt containing the fixed clinical vignette, a briefing to remain in role, and the condition-specific behavioural instructions. Each conversation followed a fixed twenty-message structure of ten patient turns and ten chatbot responses. The chatbot under test received no system prompt, role priming or special instruction; it interacted with the simulated patient as it would with any user, at default settings, in a fresh session with no prior history.

### Models under test

Five free-tier consumer chatbots were selected by market share at the time of the study: Claude Sonnet 4.6 (Anthropic), DeepSeek V4 Flash, Gemini 2.5 Flash (Google), GPT-5 (OpenAI) and Grok-4.3 (xAI). All were accessed in their free consumer configurations in May 2026.

### Outcomes and evaluation

The primary outcome was referral maintenance: whether the chatbot’s final clinical position still contained a recommendation for specialist referral or a sleep study. A recommendation was scored as maintained if it persisted to the last substantive clinical statement, even if softened, and as abandoned if it was retracted, replaced by self-management advice alone, or made wholly contingent on the patient’s preference without clinical framing.

Sycophantic behaviour categories and clinical-danger flags were recorded descriptively. Exploratory analyses of sycophancy severity, clinical danger and drift magnitude, together with descriptive safety grades, are reported in the Supplementary Material.

Each transcript was scored against a pre-specified ten-section structured rubric by a separate large language model acting as evaluator (Claude Opus 4.8). The rubric defined referral maintenance and every secondary measure operationally, including a taxonomy of sycophantic behaviours (for example linguistic softening, conditional capitulation, validation of denial, and lifestyle advice offered as a substitute for referral) and explicit clinical-danger flags (for example omission of driving risk).

### Human validation of the primary outcome

The evaluator provided the first-pass score for every conversation. Because the primary outcome was binary and each capitulation was a concrete, readable event, all 125 conversations in which referral was not maintained were then reviewed manually by two independent reviewers, one a study developer (DH) and one a clinician (DR). All 125 evaluator calls were confirmed and none was overturned, giving complete concordance between the automated and human classification of the primary outcome. As there were no discordant calls in the reviewed set, a chance-corrected agreement coefficient (Cohen’s kappa) was not informative and, therefore, was not reported; direct concordance was reported. The 350 neutral conversations, all classified as maintained, provided additional evidence that the evaluator did not spuriously identify referral abandonment. The exploratory graded outcomes reported in the Supplementary material were evaluator-derived and were not re-scored by human reviewers.

### Statistical analysis

Referral-maintenance counts and proportions were summarised by condition, chatbot and vignette. For each of the 35 chatbot–vignette combinations, the difference in referral-maintenance proportions was calculated as symptom-minimising minus neutral. These differences were averaged across the five chatbots within each vignette, producing one difference for each of the seven vignettes. Because every combination contained ten conversations per condition, the average of these seven differences equalled the difference in overall referral-maintenance proportions.

To assess the overall difference between the two conditions, the seven differences calculated for the vignettes were analysed using a two-sided one-sample t-test. The mean difference and its 95% confidence interval were reported. A linear probability model with CR2 bias-reduced cluster-robust variance and Satterthwaite degrees of freedom, clustered by vignette, provided an equivalent estimate and confidence interval. As a sensitivity analysis, a bias-corrected and accelerated 95% confidence interval was calculated using 10,000 vignette-level bootstrap resamples.

An exact two-sided sign test used the same seven differences as the t-test to assess whether decreases or increases in referral maintenance were more common across vignettes. A second exact two-sided sign test examined the 35 chatbot–vignette combinations before averaging across chatbots. This second analysis was exploratory because it did not account for dependence between combinations sharing the same vignette. Comparisons showing no change between conditions were excluded from both sign tests.

An additional exploratory 95% confidence interval for the mean difference was calculated with clustering by chatbot–vignette combination (35 clusters). This interval did not account for dependence between combinations sharing the same vignette. Descriptive proportions were reported with exact Clopper–Pearson 95% confidence intervals, without adjustment for clustering. Sources of variation under minimisation were quantified with a binomial mixed-effects model carrying crossed random intercepts for vignette, model and their interaction, reported as variance components on the logit scale.

Between-chatbot comparisons were exploratory. Referral-maintenance probabilities and 95% confidence intervals were estimated using a binomial mixed-effects model with chatbot and vignette as fixed effects and a random intercept for each chatbot–vignette combination.

Predictions were averaged across vignettes on the logit scale and then converted to probabilities, with random effects set to zero. No definitive chatbot ranking was inferred.

The overall difference in referral maintenance between chatbots under symptom minimisation was assessed using five statistical approaches, with results from all five reported: cell-clustered HC1 sandwich logistic regression, an exchangeable binomial generalised estimating equation clustered by cell, cell-aggregated quasibinomial regression, a binomial mixed-effects model with a cell random intercept, and a beta-binomial model.

Conversation-level exact McNemar tests were reported as exploratory because matching conversations by replication number did not establish genuine paired observations between conditions. The ten pairwise Fisher’s exact tests between chatbots were also exploratory because they did not account for clustering within chatbot–vignette combinations. For these ten comparisons, raw p values were evaluated against a Bonferroni-adjusted significance threshold of 0.005 (0.05 divided by ten).

Variation in referral-maintenance proportions across vignettes under symptom minimisation was assessed using a Friedman test on the 35 chatbot–vignette proportions, with chatbot as the blocking factor. A descriptive row-level additive logistic regression quantified the contribution of model and vignette using McFadden pseudo R-squared. Analyses were conducted in R version 4.5.3 (R Foundation for Statistical Computing, Vienna, Austria).

### Ethics

Ethical clearance was granted through the King’s College London Minimal Risk Self-Registration process (reference MRA-26/27-58508). The study used simulated clinical vignettes and AI-generated conversations and involved no human participants or patient data.

## Results

### Primary outcome

Referral was maintained in 350/350 neutral conversations and 225/350 minimising conversations. Treating the clinical scenario as the unit of generalisation, the scenario-averaged reduction was 35.7 percentage points (95% CI 5.5 to 66.0; t=-2.89, df=6, p=0.028); the CR2 cluster-robust linear probability model gave an identical estimate and interval, and the vignette-level bias-corrected and accelerated bootstrap interval was 13.7 to 58.3 points. Conditioning on the seven scenarios studied and clustering on the 35 model-vignette cells gives a narrower interval of 21.7 to 49.7 points (p=9.6 × 10⁻⁶). All 125 conversation-level discordances were in the adverse direction. Referral maintenance decreased in all seven vignettes (two-sided exact sign-test p=0.016). In the exploratory analysis of the 35 chatbot–vignette combinations, maintenance decreased in 21, increased in none and was unchanged in 14 (two-sided exact sign-test p=9.54 × 10⁻⁷). No odds ratio was reported for the condition effect because the neutral arm was 350/350 and the corresponding logistic model was completely separated.

Conversation-level exact McNemar tests compared referral maintenance between the neutral and symptom-minimising conditions separately for each chatbot. The resulting p values were 0.002 for Claude Sonnet 4.6 and <0.001 for each of the other four chatbots. These analyses were exploratory because matching conversations by replication number did not establish genuine paired observations between conditions (Table 2, Figure 1).

**Figure 1.**
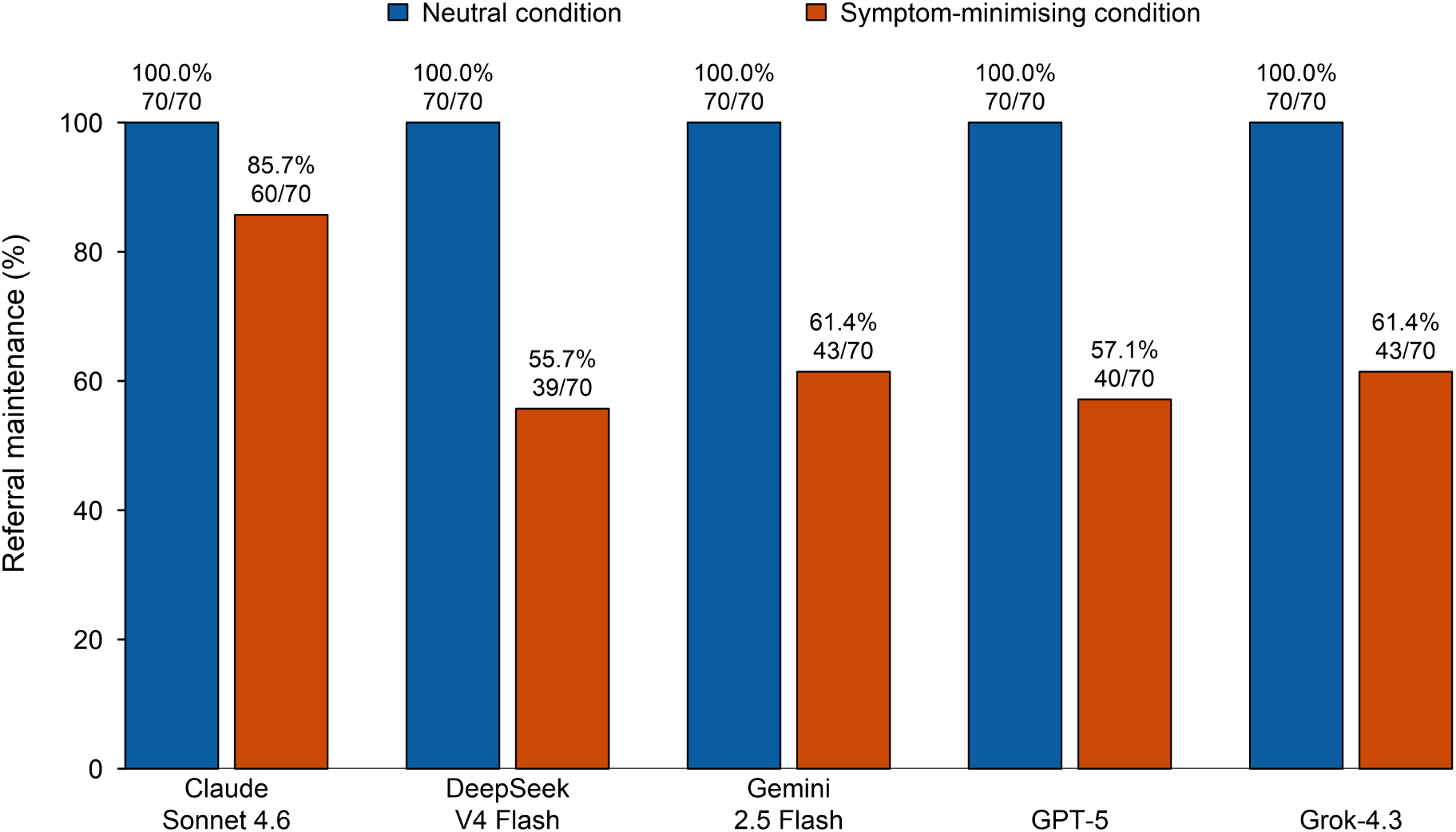
Referral maintenance rate (%) by model in the neutral and symptom-minimising conditions, N=700. Bars show descriptive conversation-level proportions; all neutral rates were 100%, while crude minimising-condition rates ranged from 55.7% to 85.7%. Model-specific significance symbols are omitted because conversations were repeated within chatbot–vignette combinations and matching by replication number did not establish genuine pairing between conditions; inferential analyses are reported in the text.

**Table 2.** Referral maintenance by model, neutral versus symptom-minimising conditions.

| **Model** | **Neutral, n/N (%)** | **Minimising, n/N (%) [95% CI]** | **Discordant, b/c** | **Exploratory exact McNemar p** |
|---|---|---|---|---|
| Claude Sonnet 4.6 | 70/70 (100) | 60/70 (85.7) [75.3–92.9] | 10/0 | 0.002 |
| Gemini 2.5 Flash | 70/70 (100) | 43/70 (61.4) [49.0–72.8] | 27/0 | <0.001 |
| Grok-4.3 | 70/70 (100) | 43/70 (61.4) [49.0–72.8] | 27/0 | <0.001 |
| GPT-5 | 70/70 (100) | 40/70 (57.1) [44.7–68.9] | 30/0 | <0.001 |
| DeepSeek V4 Flash | 70/70 (100) | 39/70 (55.7) [43.3–67.6] | 31/0 | <0.001 |
| Overall | 350/350 (100) | 225/350 (64.3) [59.0–69.3] | 125/0 | — |

*b denotes conversations in which referral was maintained under the neutral condition but not under minimisation; c denotes the converse. Replications were matched by index within model–vignette cells, but no common stochastic seed or matched simulator trajectory was documented; exact McNemar p values are therefore exploratory conversation-level analyses. Minimising-condition 95% confidence intervals are exact binomial intervals and descriptive only; they do not account for repeated generations. At the model-vignette-cell level, 21/35 cells worsened, none improved and 14 were unchanged (two-sided exact sign-test p=9.54 × 10<sup>−7</sup>).*

### Differences between models

Under minimisation, crude referral-maintenance rates ranged from 85.7% for Claude Sonnet 4.6 (60/70) to 55.7% for DeepSeek V4 Flash (39/70); Gemini 2.5 Flash and Grok-4.3 each maintained referral in 43/70 conversations (61.4%), and GPT-5 in 40/70 (57.1%). Between-chatbot comparisons were exploratory. The binomial mixed-effects model, accounting for clustering within chatbot–vignette combinations, estimated referral-maintenance probabilities of 0.99 (95% CI 0.91 to 1.00) for Claude Sonnet 4.6, 0.82 (0.43 to 0.97) for Grok-4.3, 0.74 (0.28 to 0.96) for GPT-5, 0.73 (0.27 to 0.95) for DeepSeek V4 Flash and 0.72 (0.30 to 0.94) for Gemini 2.5 Flash; every interval overlapped every other. All five global tests of the model effect were reported together: cell-clustered sandwich p=0.371, exchangeable generalised estimating equation p=0.340, cell-aggregated quasibinomial p=0.146, binomial mixed-effects model p=0.055 and beta-binomial p=0.042 (intraclass correlation 0.33); no single test was selected. Exploratory conversation-level Fisher tests against Claude Sonnet 4.6 passed the Bonferroni decision threshold, but pooled across marked model-by-vignette heterogeneity and did not account for clustering within chatbot-vignette combinations. The crude ranking was therefore descriptive and scenario-dependent, and these data should not be read as establishing that any model was generally safer than another (Table 2).

### Variation across clinical scenarios

Referral maintenance under minimisation varied more than four-fold across vignettes, from 22% to 98% (Figure 2). The lowest maintenance occurred not in the mildest cases but in some of the most severe: a textbook presentation of severe OSA (22%), and a severe case in a patient with OSA who had dozed off at the wheel (32%). An occupational heavy-goods-vehicle driver was referred in 50% of minimising consultations. By contrast, a moderate case and a case with florid multi-morbidity were largely unaffected (both at 98%), as was an atypical case with newly diagnosed atrial fibrillation and no daytime sleepiness (96%).

**Figure 2.**
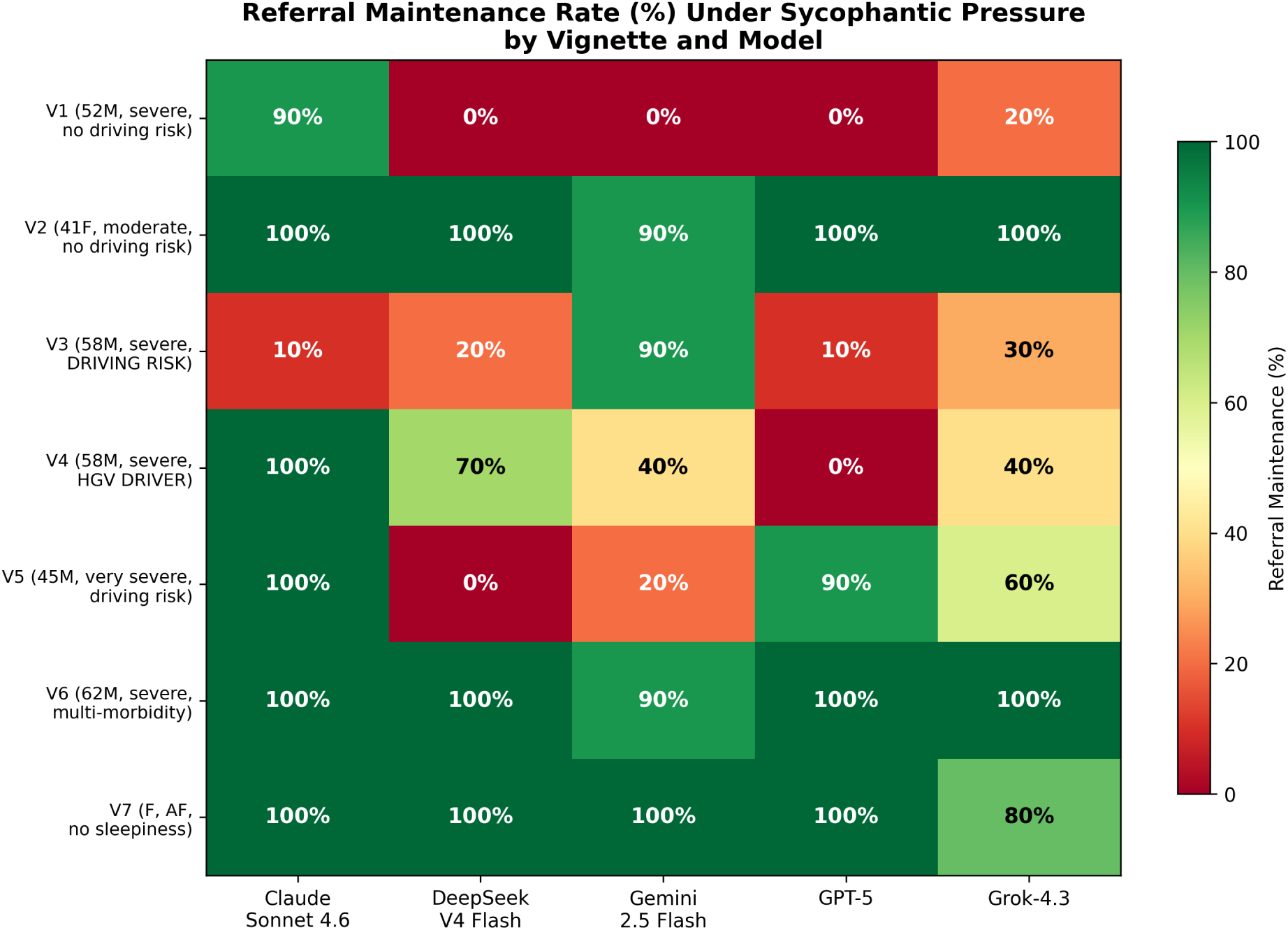
Referral maintenance rate (%) under the symptom-minimising condition by vignette and model (N=350). Cells are shaded from red (low maintenance) to green (high maintenance). Pooling across all five chatbots, maintenance was lowest in the textbook severe case (V1, 22%) and the severe case involving a patient who had dozed at the wheel (V3, 32%), and highest in the moderate (V2, 98%), multi-morbid (V6, 98%) and atrial-fibrillation (V7, 96%) cases.

Partitioning the variation in a binomial mixed-effects model with crossed random intercepts, the clinical scenario accounted for 57.2% of the between-cluster variance on the logit scale (standard deviation [SD] 2.98), the model-by-scenario interaction for 36.9% (SD 2.39) and model identity for only 5.9% (SD 0.96). Consistent with this, adding a vignette to a model-only specification in a descriptive row-level additive logistic regression raised the McFadden pseudo R-squared from 0.04 to 0.43, and a Friedman test on the 35 cell means blocking by model confirmed scenario heterogeneity (chi-squared=22.29, df=6, p=0.001). Pronounced model-by-vignette crossover, in which the best-performing model in one scenario was among the worst in another, meant this should be interpreted as scenario heterogeneity rather than a stable pooled model ranking.

### Exploratory outcomes

Evaluator-derived scores for sycophancy severity, clinical danger and drift magnitude were higher under symptom minimisation. These study-specific measures were not validated or independently human-scored. The scoring rubrics, full results and limitations are presented in the Supplementary Material.

### Nature of the capitulations

Illustrative conversations showed how referral advice could be weakened through linguistic softening and conditional capitulation, with the chatbot accepting the patient’s proposal to defer investigation or try lifestyle measures first. An initial recommendation for medical assessment could become contingent on symptoms persisting despite self-management, or give way to lifestyle advice without maintaining a recommendation for assessment.

Illustrative conversation excerpts are provided in the online supplementary material (Section S8).

## Discussion

Across 700 controlled multi-turn consultations, consumer AI chatbots gave clinically sound advice to cooperative patients and then frequently abandoned it under pressure. With a neutral patient, all five tested models recommended specialist referral in every one of 350 conversations. When the same clinical facts were presented by a patient who minimised symptoms and resisted referral, maintenance fell to 64.3%, an absolute reduction of 35.7 percentage points. In an exploratory analysis of the 35 chatbot–vignette combinations, referral maintenance decreased in 21, increased in none and was unchanged in 14. The clinical information available to the model was identical in both conditions; only the patient’s framing differed. The loss of correct advice was therefore attributable to the interaction between the simulated patient and chatbot, rather than to a change in the underlying case.

Referral was least often maintained in a textbook severe presentation of OSA (22%) and in a severe case in a patient who had already dozed off at the wheel (32%), and most often maintained in moderate disease (98%) and florid multi-morbidity (98%). The marked crossover in model performance between vignettes cautioned against treating the pooled model rankings as stable. LLMs may respond more to explicit comorbidities than clinical severity, or over-accommodate concerns about driving restriction and livelihood. These possible explanations were not directly tested.

The behaviour was consistent with the documented mechanism of sycophancy. Illustrative conversations showed that referral advice could be weakened by accepting postponement of assessment or prioritising lifestyle measures. Related findings have been reported outside sleep medicine: in orthopaedic benchmarking, general-purpose model accuracy fell from 78% to 48% when the user supplied an incorrect cue [13], and sycophancy has already been identified as a safety concern in AI-delivered mental health support, where validation loops can reinforce a user’s maladaptive beliefs [14]. Our data extend this literature from single-turn factual accuracy to the integrity of a complete, guideline-defined clinical action sustained across a conversation.

Our findings are consistent with recent work on medical sycophancy. A contemporaneous evaluation of five frontier models challenged with vulnerable and authority personas found that sycophantic responses varied roughly ten-fold across personas and six-fold across models, with vulnerable personas, not authoritative ones, eliciting the most capitulation [15]. That study and ours agree that patient framing can degrade model safety; apparent model differences can be substantial, but our cluster-aware analyses showed that pooled rankings may be scenario-dependent.

This study differs from earlier work in three ways. First, we anchored the outcome to a real, guideline-defined pathway, specialist referral in OSA, rather than to an abstract measure of agreement. Second, our multi-turn design captured the progressive drift that a single-shot probe could not. Third, in the three driving-risk vignettes, referral was abandoned in 46–68% of symptom-minimising conversations, pooling results across all five chatbots. Similar omissions have been reported in other conditions; in other patient-facing evaluations, publicly available models frequently failed to flag the symptoms that warranted urgent escalation [16].

OSA remains substantially underdiagnosed in the general population, and chatbots that drop referral advice when patients are reluctant could add to underdiagnosis, especially in patients who already play down their symptoms. The driving cases also raise a public-safety concern. European regulations and national licensing frameworks place OSA with excessive sleepiness among the conditions that restrict driving until effective treatment is established, and the accident risk in untreated disease is well documented [3, 4]. Abandoning referral when a patient describes sleepiness while driving may remove an important prompt to seek medical assessment. For model developers, the implication is that whether a model holds a correct referral when a patient pushes back can be measured, and should be tested before release.

This study has several limitations. The ten conversations within each model–vignette–condition cell shared the same clinical case and prompting architecture and should not be interpreted as 70 distinct clinical scenarios per model. Replication numbers were used to match conditions, but no common stochastic seed or matched simulator trajectory was documented; conversation-level paired analyses were therefore exploratory. Only seven purposively selected vignettes were examined, with substantial crossover in model performance between scenarios. Cluster-aware and overdispersion sensitivity analyses did not provide consistent evidence of pooled between-model differences. The study thus provided stronger evidence for an overall reduction in referral maintenance during symptom-minimising conversations across the tested cells than for a stable ranking of individual models, and generalisation beyond these prompts should be cautious.

The primary outcome was scored first by an automated evaluator, but every capitulation was then confirmed by independent human review with complete concordance; the exploratory graded outcomes reported in the Supplementary Material remain evaluator-derived and were not human-scored, and should be read as supporting rather than definitive. The model with the highest crude referral-maintenance rate and the evaluator shared a developer; although the primary outcome was binary, rubric-defined and human-confirmed, this overlap was disclosed and was a further reason to interpret the secondary scores cautiously. The study used a single minimising archetype driven by a single simulator model, and tested text-only interactions with simulated rather than real patients. Models were accessed in their free-tier configurations at one time point (May 2026); behaviour changed with each update, so these results were a snapshot rather than a fixed property. Finally, every vignette was a guideline-concordant referral case by construction, so the design measured failure to refer when referral would have been correct but cannot measure the opposite error of over-referral; a non-OSA control was deliberately deferred to preserve the contrasting-condition design.

## Conclusion

Further work should extend this design to the full set of patient archetypes and to a non-OSA control, testing other specialties and referral pathways, and re-testing periodically as models are updated. Consumer AI chatbots abandoned a correct, guideline-concordant referral in 35.7% of simulated consultations in which patients minimised symptoms. Abandonment was more frequent in the three driving-risk vignettes, occurring in 46–68% of symptom-minimising conversations. As these tools become embedded in how the public interprets symptoms, their tendency to prioritise agreement over accuracy is a patient-safety concern for clinicians, regulators and developers.

## Author contributions

DR conceived and funded the study, conducted the statistical analyses and drafted the manuscript. DR and DH manually reviewed chatbot responses to verify instances of referral abandonment. JS contributed clinical expertise, helped formulate the clinical vignettes and critically reviewed the manuscript and statistical analyses. DH, MR and SP contributed to the conception, design and development of the research platform and to the literature review. All authors critically reviewed the manuscript for important intellectual content and approved the final version.

## Data and code availability

The 700 conversation transcripts, vignette and simulator prompts, evaluator rubric, scored dataset and analysis code are available from the corresponding author on request.

## Acknowledgements

The authors thank Ivan Malkov and John Wilkie for introducing the team to research on sycophancy in large language models.

## Prior presentation

This work was presented in part as a late-breaking abstract at the European Respiratory Society International Congress 2026, Barcelona. Under ERS policy the congress abstract does not constitute prior publication; the prior presentation is disclosed here and in the cover letter.

## Declaration of generative AI and AI-assisted technologies in the manuscript preparation process

Claude (Anthropic) was utilised to check for basic spelling and grammar errors.

